# Aerosolized Ad5-nCoV booster vaccination elicited potent immune response against the SARS-CoV-2 Omicron variant after inactivated COVID-19 vaccine priming

**DOI:** 10.1101/2022.03.08.22271816

**Authors:** Zhe Zhang, Shipo Wu, Yawei Liu, Kailiang Li, Pengfei Fan, Xiaohong Song, Yudong Wang, Zhenghao Zhao, Xianwei Zhang, Jin Shang, Jinlong Zhang, Jinghan Xu, Yao Li, Yaohui Li, Jipeng Zhang, Kefan Fu, Busen Wang, Meng Hao, Guanying Zhang, Pengwei Long, Ziyu Qiu, Tao Zhu, Shuling Liu, Yue Zhang, Fangze Shao, Peng Lv, Yilong Yang, Xiaofan Zhao, Yufa Sun, Lihua Hou, Wei Chen

## Abstract

The SARS-CoV-2 Omicron variant has become the dominant SARS-CoV-2 variant around the world and exhibits immune escape to current COVID-19 vaccines to some extent due to its numerous spike mutations. Here, we evaluated the immune responses to booster vaccination with intramuscular adenovirus-vectored vaccine (Ad5-nCoV), aerosolized Ad5-nCoV, a recombinant protein subunit vaccine (ZF2001) or homologous inactivated vaccine (CoronaVac) in those who received two doses of inactivated COVID-19 vaccines 6 months prior. We found that the Ad5-nCoV booster induced potent neutralizing activity against the wild-type virus and Omicron variant, while aerosolized Ad5-nCoV generated the greatest neutralizing antibody responses against the Omicron variant at day 28 after booster vaccination, at 14.1-fold that of CoronaVac, 5.6-fold that of ZF2001 and 2.0-fold that of intramuscular Ad5-nCoV. Similarly, the aerosolized Ad5-nCoV booster produced the greatest IFNγ T-cell response at day 14 after booster vaccination. The IFNγ T-cell response to aerosolized Ad5-nCoV was 12.8-fold for CoronaVac, 16.5-fold for ZF2001, and 5.0-fold for intramuscular Ad5-nCoV. Aerosolized Ad5-nCoV booster also produced the greatest spike-specific B cell response. Our findings suggest that inactivated vaccine recipients should consider adenovirus-vectored vaccine boosters in China and that aerosolized Ad5-nCoV may provide a more efficient alternative in response to the spread of the Omicron variant.

## Introduction

More than 5.9 million people have died from COVID-19 worldwide since the start of the pandemic*(1)*. The COVID-19 vaccines studied to date are highly effective against severe disease and death. However, immunity from the COVID-19 vaccines is waning, and variants capable of different degrees of immune evasion are continuously emerging; thus, there is a clear and urgent need for booster vaccination to increase vaccine effectiveness against severe disease and death.

More than 100 countries worldwide have already issued recommendations on booster or additional vaccination*(2)*. Both homologous and heterologous booster regimens including mRNA vaccines, adenovirus-vectored vaccines, inactivated vaccines and recombinant protein vaccines are immunologically effective, and no safety issues have been observed. In Israel, a booster dose of the BNT162b2 vaccine induced a more than 10-fold decrease in the relative risk of confirmed infection and severe illness compared with that of the nonbooster group*(3)*. The effectiveness of a booster dose of the BNT162b2 vaccine reached 93% for admission to the hospital compared with that upon receipt of only two doses at least 5 months prior*(4)*.

In China, seven COVID-19 vaccines have been authorized for use, including five inactivated vaccines, an adenovirus-vectored vaccine (Ad5-nCoV, Convidecia) and a recombinant protein subunit vaccine (ZF2001, Zifivax). These vaccines have been shown to be efficacious in preventing mild to severe COVID-19*(5-7)*. To date, more than 3.0 billion doses of COVID-19 vaccines have been administered in China, and over 95% of these doses were of the inactivated vaccines. Heterologous booster vaccination was recently approved, and more than 460 million individuals have received homologous booster vaccination in China*(8)*.

Booster vaccination strategies based on inactivated vaccine priming have been well studied, and heterologous vaccination regimens induce immune responses that are superior to those induced by homologous regimens*(9-12)*. Clemens et al. reported that the increases in specific IgG titers from baseline to 28 days were 12-fold for CoronaVac (the inactivated vaccine), 152-fold for BNT162b2, 90-fold for ChAdOx1 nCoV-19, and 77-fold for Ad26.COV2.S after booster vaccination in CoronaVac-primed recipients*(11)*. Li et al. reported that neutralizing antibody titers were increased by 79-fold for Ad5-nCoV booster vaccination and by 15-fold for CoronaVac booster vaccination from before booster vaccination to day 14 after booster vaccination in subjects who received two doses of CoronaVac*(12)*.

To optimize the booster vaccination regimen in persons who have received two doses of inactivated vaccines, we performed a head-to-head immunological comparison of intramuscular Ad5-nCoV, aerosolized Ad5-nCoV, a recombinant protein subunit vaccine (ZF2001) and homologous CoronaVac booster administration in inactivated vaccine-primed recipients who were vaccinated 6 months prior.

## Results

### Baseline characteristics of the participants

In this study, 904 subjects who received two doses of inactivated vaccine 6 months prior were assigned to 4 groups for booster vaccination. The participants in two of the groups received Ad5-nCoV booster vaccination by different delivery routes. A total of 229 participants in the Ad5-nCoV-IM group were intramuscularly vaccinated with 5×10^10^ viral particles of Ad5-nCoV per dose, and 223 participants in the Ad5-nCoV-IH group were vaccinated with 1×10^10^ viral particles of aerosolized Ad5-nCoV per dose. A total of 219 participants in the ZF2001 group received the recombinant protein subunit vaccine (ZF2001), and 233 participants in the CoronaVac group received an inactivated vaccine (CoronaVac). The baselines of the participants, including age, sex, interval to prime vaccination, and anti-SARS-CoV-2 neutralizing antibodies, were comparable among the four group.

### Binding antibody responses

Concentrations of anti-RBD IgG antibodies were assessed at baseline and at 7, 14 and 28 days after booster vaccination (Fig. 1). In all groups, IgG antibody concentrations peaked at day 14 and dropped slightly at day 28 after booster vaccination. Intramuscular injection of Ad5-nCoV elicited the most significant and rapid increase in anti-RBD IgG antibodies by 30-fold compared to baseline, followed by CoronaVac, with a 9-fold increase compared to baseline at 7 days (Fig. 1A). ZF2001 booster vaccination slightly enhanced anti-RBD IgG antibodies, with a 3-fold increase compared to baseline, whereas the administration of aerosolized Ad5-nCoV did not alter the anti-RBD IgG antibody levels. The seroconversion rate (at least a fourfold increase in postvaccination titer from baseline) reached 93.4% for Ad5-nCoV-IM and 79.3% for CoronaVac (Fig. 1B). After 14 days, the magnitude of the IgG response was sharply enhanced, especially in the heterologous booster groups (Fig. 1C). The median fold increase from baseline to day 14 was 464 (Interquartile Range (IQR), 210-1097) for Ad5-nCoV-IM, 523 (IQR, 137-1336) for Ad5-nCoV-IH, 174 (IQR, 58-488) for ZF2001 and 61 (IQR, 30-124) for CoronaVac. All heterologous regimens were superior to homologous CoronaVac booster vaccination (P<0.0001), and both intramuscular and aerosolized Ad5-nCoV vaccination induced similarly increases in the IgG response compared to that of ZF2001 (P<0.0001). At day 28, the median fold increase from baseline was 281 (IQR, 115-507) for Ad5-nCoV-IM, 361 (IQR, 149-841) for Ad5-nCoV-IH, 120 (IQR, 42-360) for ZF2001 and 47 (IQR, 22-108) for CoronaVac. The geometric mean concentration (GMC) of IgG in the Ad5-nCoV-IH group was higher than those in the ZF2001 and CoronaVac groups (P<0.0001), and similar to that in the Ad5-nCoV-IM group (P=0.2513). These data suggest that heterologous boosting with Ad5-nCoV via different routes can elicit significantly higher RBD-specific IgG levels than ZF2001 or CoronaVac.

**Fig. 1.**
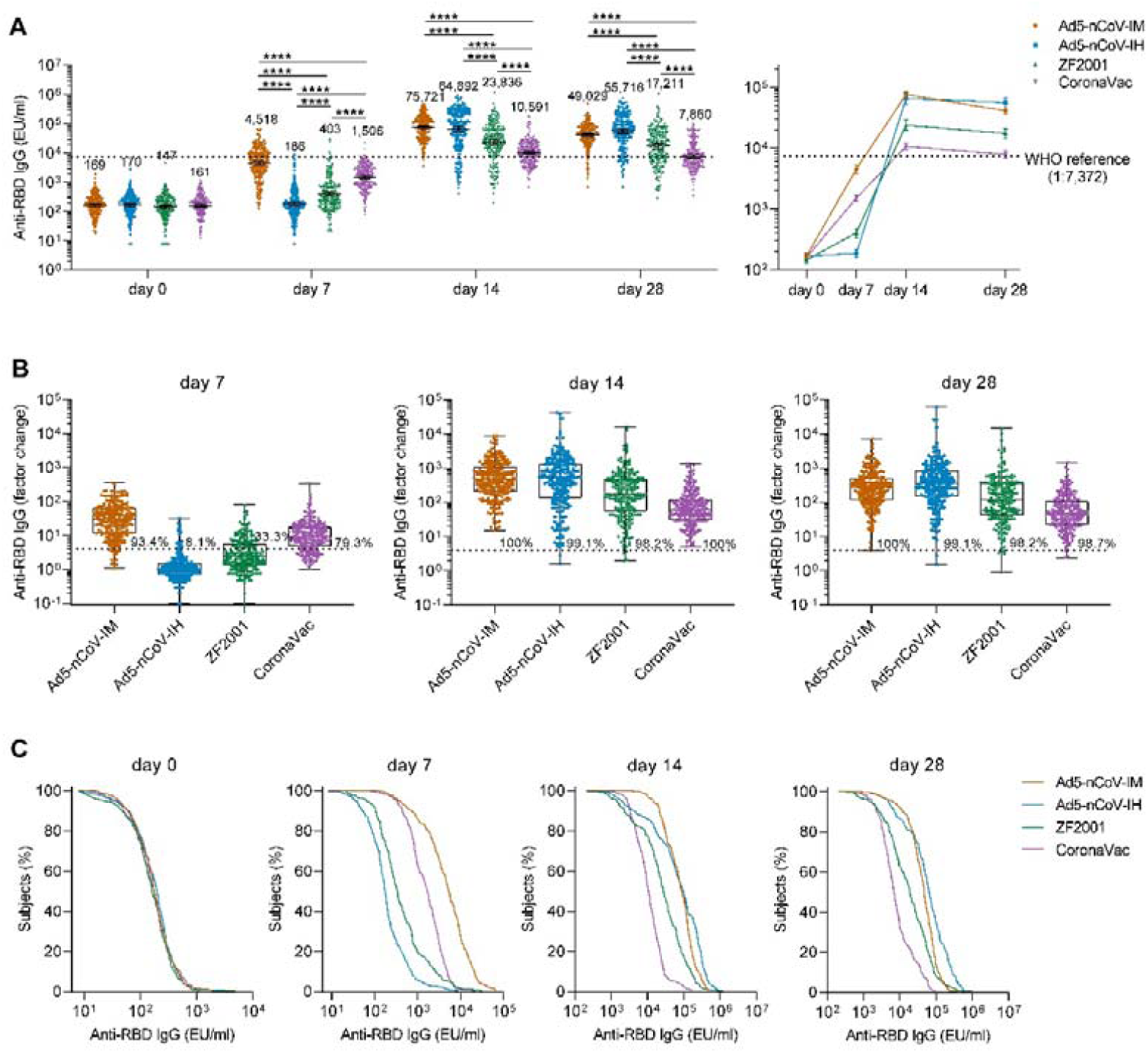
SARS-CoV-2 RBD-specific binding antibody responses. (A) GMCs of SARS-CoV-2 RBD-specific IgG antibodies at day 0 (before booster vaccination) and days 7, 14 and 28 after booster vaccination in the four groups. Error bars indicate 95% CIs, the numbers above the bars are GMCs for the group, and connecting lines reflect geometric means. The WHO reference (1,000 binding antibody units (BAU) ml^-1^ in serum) is equivalent to an RBD-specific IgG antibody titer of 1:7,372. Statistical significance was determined by Kruskal–Wallis ANOVA with Dunn’s multiple comparisons tests. **** P<0.0001. (B) Per-participant factor changes that were calculated by dividing the after-booster response by the before-booster response for RBD-specific binding antibodies. Data are presented in box-and-whisker plots. The whiskers indicate the range, the top and bottom of the boxes indicate the interquartile range, and the horizontal line within each box indicates the median. The dashed line indicates a factor change of 4 (the lower limit of seroconversion), and the number above the dashed line indicates the seroconversion of RBD-specific IgG responses. (C) RBD-specific IgG reverse cumulative distribution curves for each group at day 0, 14 and 28 after the booster. Reverse cumulative distribution curves denote the percentage of participants that reach different level of antibody concentration.

### Neutralizing antibody responses against wild-type SARS-CoV-2 and the Omicron variant

Approximately 50 samples in each group were evaluated for neutralizing antibody responses using pseudovirus-based neutralization assays (Fig. 2). Before booster vaccination, only 6.3%∼11.8% of participants had a weak pseudovirus neutralization antibody (PNAb) titer. Generation of PNAb against wild-type SARS-CoV-2 was significantly increased after booster vaccination in all groups (Fig. 2A). At day 14, participants who received intramuscular Ad5-nCoV had the highest geometric mean titer (GMT) of PNAb at 970 (95% CI = 727-1294), compared with a GMT of 567 (95% CI = 341-944) in the Ad5-nCoV-IH group (P=0.4634), a GMT of 308 (95% CI = 204-466) in the ZF2001 group (P=0.0007) and a GMT of 139 (95% CI = 107-181) in the CoronaVac group (P<0.0001) (Fig. 2A). An increased PNAb response was also observed when heterologous aerosolized Ad5-nCoV (P<0.0001) or ZF2001 (P=0.0101) was compared with the homologous CoronaVac. At day 28, the PNAb level in the Ad5-nCoV-IH group slightly differed from its IgG response, as an increased GMT of 874 (95% CI = 569-1342) was observed, while those in the Ad5-nCoV-IM, ZF2001 and CoronaVac groups decreased to 628 (95% CI = 455-868), 210 (95% CI = 137-321) and 69 (95% CI = 51-93), respectively.

**Fig. 2.**
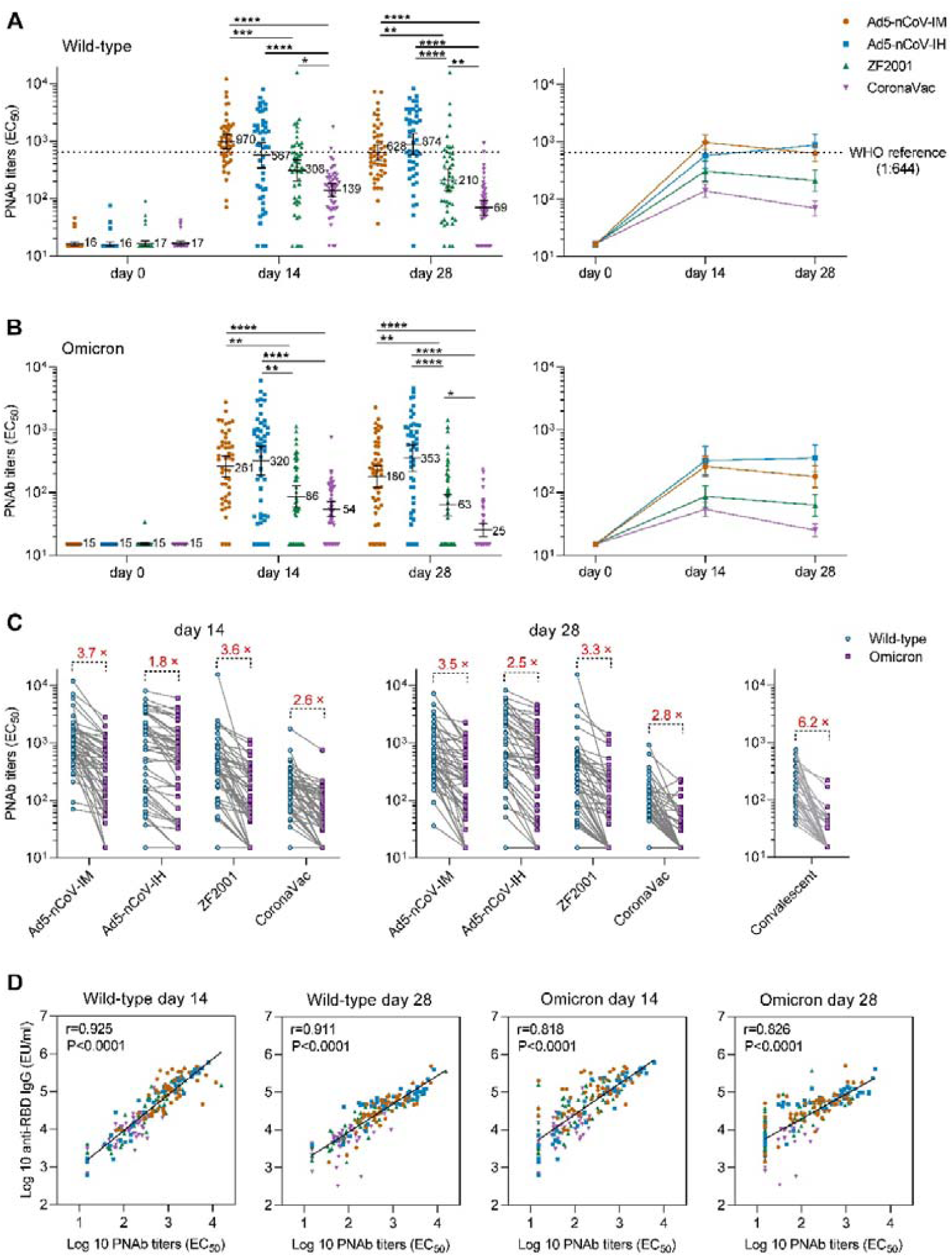
SARS-CoV-2 PNAb responses. (A and B) GMTs of SARS-CoV-2 PNAb to wild-type SARS-CoV-2 (A) or the Omicron variant (B) at day 0 (before booster vaccination) and 14 and 28 days after booster vaccination in the four groups. Error bars indicate 95% CIs, the numbers on the right of the bars are GMTs for the group, and connecting lines reflect geometric means. The WHO reference (1,000 binding antibody units (BAU) ml^-1^ in serum) is equivalent to a PNAb titer of 1:644 to the wild-type and lower than the limit of detection to the Omicron variant of SARS-CoV-2. Statistical significance was determined by Kruskal–Wallis ANOVA with Dunn’s multiple comparisons tests. * P<0.05, ** P<0.01, *** P<0.001, **** P<0.0001. (C) Comparison of PNAb titers between wild-type SARS-CoV-2 and the Omicron SARS-CoV-2 variant in four groups at days 14 and 28 after booster vaccination. Sera from a group of 29 convalescent patients infected with wild-type SARS-CoV-2 in Wuhan who had recovered for half a year were also included. The numbers on the top are the fold decline in PNAb GMTs from the wild-type to the Omicron variant of SARS-CoV-2 in each group. (D) Correlation between PNAb and RBD-specific IgG antibodies. Spearman’s correlation and linear regression (diagonal lines) analyses were performed with log-transformed data. Spearman r and corresponding two-tailed *P* values are shown in the top left corner.

Similar kinetics were observed for the PNAb response against the Omicron variant. Only one participant had positive neutralizing antibody responses to the Omicron variant before booster vaccination. Both the Ad5-nCoV-IM and Ad5-nCoV-IH groups exhibited a remarkably higher PNAb level than the other two groups at days 14 and 28 (Fig. 2B). Specifically, participants in the Ad5-nCoV-IH group rapidly generated the most robust response at day 14, with a GMT of 320 (95% CI = 191-538), followed by the Ad5-nCoV-IM group, with a GMT of 261 (95% CI = 178-382); the ZF2001 group (P=0.001), with a GMT of 86 (95% CI = 59-127); and the CoronaVac group (P<0.0001), with a GMT of 54 (95% CI = 42-71). Prominent cross-neutralization potential was observed in the Ad5-nCoV-IH group, with the lowest fold reduction in the levels of PNAb against wild-type SARS-CoV-2 compared to the Omicron variant at days 14 (1.8 ×) and 28 (2.5 ×) after booster vaccination, while there was a 6.2-fold decrease in the convalescents at approximately 6 months postinfection (Fig. 2C). PNAb responses were substantially correlated with IgG levels after booster vaccination in all groups, regardless of the variant, especially in the Ad5-nCoV-IH group (Fig. 2D). These results demonstrate that heterologous booster vaccination with Ad5-nCoV or ZF2001 provides a higher PNAb response against SARS-CoV-2 and the Omicron variant than that provided by homologous booster vaccination with CoronaVac.

### Spike-specific IgG B-cell responses

To further investigate the ability of the boosters to activate B cells to produce specific antibodies, spike-specific IgG spots were detected at baseline and at 14 and 28 days after booster vaccination in R848-activated peripheral blood mononuclear cells (PBMCs) from approximately 50 participants in each group (Fig. 3). Significantly more spike-specific IgG spots were detected in all groups after booster vaccination, with the peak level measured at day 14 (Fig. 3A and C). The median number of IgG spots per 10^6^ PBMCs at day 14 was 600 (IQR, 258-1350) for Ad5-nCoV-IM, 1250 (IQR, 380-3830) for Ad5-nCoV-IH, 100 (IQR, 30-440) for ZF2001 and 80 (IQR, 30-300) for CoronaVac; these values changed to 265 (IQR, 63-658), 360 (IQR, 155-730), 70 (IQR, 30-200) and 90 (IQR, 15-165) at day 28. At day 14, the Ad5-nCoV vaccine induced a significant increase in the spike-specific IgG spot response in the Ad5-nCoV-IM group versus either the ZF2001 group (P=0.0005) or the CoronaVac group (P<0.0001) and the Ad5-nCoV-IH group versus either the ZF2001 group (P<0.0001) or the CoronaVac group (P<0.0001). A total of 82.6% (95% CI, 68.6%-92.2%) and 60.9% (95% CI, 45.4%-74.9%) of participants in the Ad5-nCoV-IM group and 89.4% (95% CI, 76.9%-96.5%) and 71.7% (95% CI, 56.5%-84.0%) of participants in the Ad5-nCoV-IH group exhibited a 4-fold or more increase in the median number of spike-specific IgG spots at days 14 and 28, respectively, which was significantly higher than that of the ZF2001 and CoronaVac groups (Fig. 3B). The RBD-specific IgG concentrations in the culture supernatant of R848-actived PBMCs was consistent with the spike-specific IgG spot response among the groups, except for the CoronaVac group, which showed a lower RBD IgG response than the ZF2001 group (Fig. 3D). A positive correlation between spike IgG spots and RBD IgG concentrations in the culture was found at days 14 and 28 (Fig. 3E). Considering the RBD-IgG and neutralizing antibody response, we conclude that booster vaccination with Ad5-nCoV, especially aerosolized Ad5-nCoV, substantially improved the immune response in inactivated vaccine-primed recipients.

**Fig. 3.**
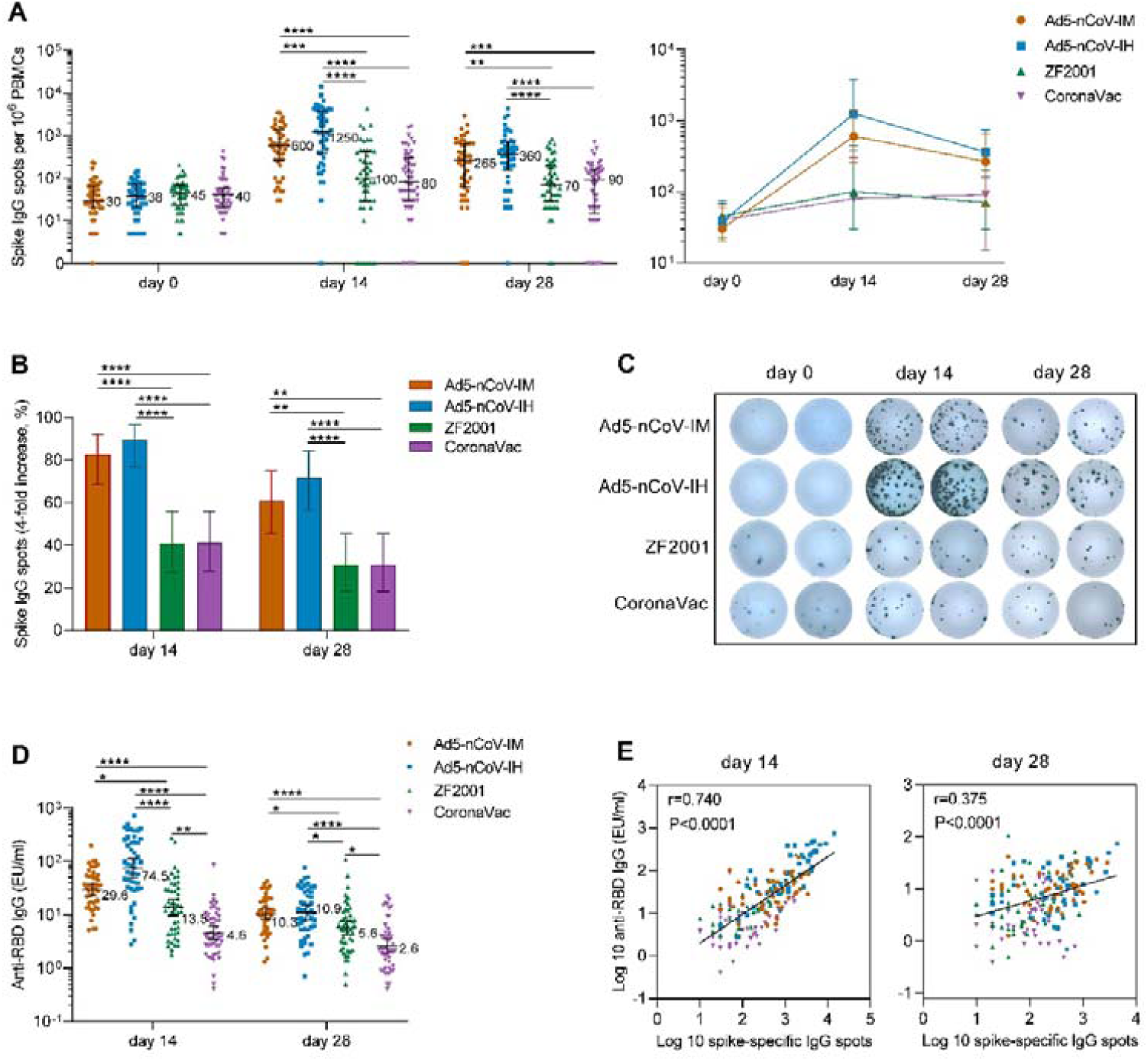
SARS-CoV-2 spike-specific IgG B-cell responses. (A) Median numbers of SARS-CoV-2 spike-specific IgG spots at day 0 (before booster vaccination) and days 14 and 28 after booster vaccination in the four groups. Error bars indicate IQRs, the numbers on the right of the bars are medians for the group, and connecting lines reflect medians. Statistical significance was determined by Kruskal–Wallis ANOVA with Dunn’s multiple comparisons tests. ** P<0.01, *** P<0.001, **** P<0.0001. (B) Percentage of participants with a fourfold increase in spike-specific IgG spots. Error bars indicate 95% CIs. Statistical significance was determined by two-sided χ^2^ tests or Fisher’s exact test, ** P<0.01, *** P<0.001, **** P<0.0001. (C) Representative spike-specific IgG spots at days 0, 14 and 28 after booster vaccination in the four groups. (D) GMTs of SARS-CoV-2 RBD-specific IgG antibodies in the culture supernatant of R848- and IL-2-activated PBMCs at days 14 and 28 after booster vaccination. Error bars indicate 95% CIs, and the numbers on the right of the bars are GMTs for the group. Statistical significance was determined by Kruskal–Wallis ANOVA with Dunn’s multiple comparisons tests. * P<0.05, ** P<0.01, **** P<0.0001. (E) Correlation between spike-specific IgG spots and RBD-specific IgG antibodies in the supernatant of R848- and IL-2-activated PBMCs. Spearman’s correlation and linear regression (diagonal lines) analyses were performed with log-transformed data. Spearman r and the corresponding two-tailed *P* values are shown in the top left corner.

### Spike-specific IFNγ responses

Spike-specific IFNγ responses were detected at baseline and at 14 and 28 days after booster vaccination to determine the T-cell response in approximately 50 participants of each group (Fig. 4). Booster vaccinations induced a rapid spike-specific IFNγ response compared with baseline levels. The participants who received the Ad5-nCoV booster vaccination showed higher T-cell responses than those who received ZF2001 or CoronaVac, and aerosolized Ad5-nCoV booster vaccination induced the greatest IFNγ response (Fig. 4A). The response was 100% (95% CI, 92.6%-100.0%) and 95.7% (95% CI, 85.2%-99.5%) with the aerosol Ad5-nCoV booster and 85.4% (95% CI, 72.2%-93.9%) and 68.8% (95% CI, 53.7%-81.3%) with the intramuscular Ad5-nCoV booster at days 14 and 28, respectively; both performed better than ZF2001 and CoronaVac (response, <25% for ZF2001 and <42% for CoronaVac) (Fig. 4B). Spike-specific IFNγ responses were positively correlated with the presence of spike-specific IgG spots (Fig. 4C).

**Fig. 4.**
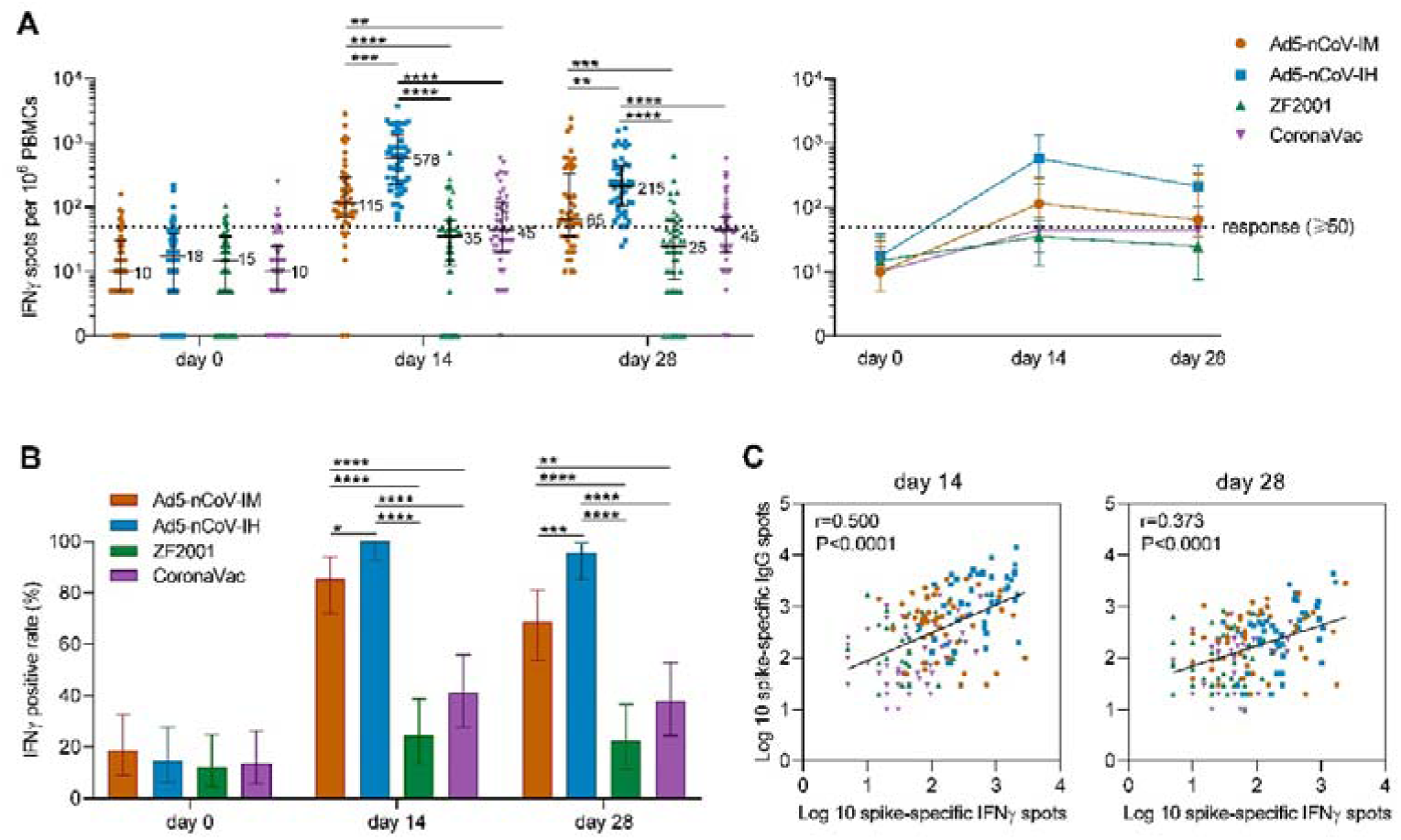
SARS-CoV-2 spike-specific IFNγ ELISpot responses. (A) Median numbers of SARS-CoV-2 spike-specific IFNγ spots at day 0 (before booster vaccination) and days 14 and 28 after booster vaccination in the four groups. Error bars indicate IQRs, the numbers on the right of the bars are medians for the group, and connecting lines reflect medians. The dashed line indicates the lower limit of the positive response (50 spots per 10^6^ PBMCs). Statistical significance was determined by Kruskal–Wallis ANOVA with Dunn’s multiple comparisons tests. ** P<0.05, ** P<0.01, *** P<0.001, **** P<0.0001. (B) Percentage of participants with a positive IFNγ response at days 0, 14 and 28 after booster vaccination. Responses were considered positive if there were ≥50 spike-specific spot-forming cells (SFCs) per 10^6^ PBMCs and the ratio of spots in the stimulated wells to spots in background wells was no less than 2.1. Error bars indicate 95% CIs. Statistical significance was determined by two-sided χ^2^ tests or Fisher’s exact test, ** P<0.01, *** P<0.001, **** P<0.0001. (C) Correlation between spike-specific IFNγ spots and spike-specific IgG spots at days 14 and 28 after booster vaccination. Spearman’s correlation and linear regression (diagonal lines) analyses were performed with log-transformed data. Spearman r and the corresponding two-tailed *P* values are shown in the top left corner.

## Discussion

In China, where more than 90% of individuals vaccinated against COVID-19 received inactivated vaccines, we evaluated the immunogenicity of homologous and heterologous boosters in adults who received prime vaccination with two doses of the inactivated COVID-19 vaccine approximately 6 months prior. Both homologous and heterologous booster vaccination led to an increase in levels of spike RBD-specific binding antibodies, neutralizing antibodies, the B-cell response and T-cell responses from day 14 after booster vaccination, but these increases were highest in participants who received heterologous regimens with an adenovirus-based COVID-19 vaccine.

Booster vaccination with the Ad5-nCoV vaccine induced a superior T-cell response and neutralizing antibody responses compared to those induced by the homologous inactivated vaccine booster or heterologous recombinant protein vaccine booster, regardless of whether intramuscular injection or aerosol inhalation was used*(13)*. At day 7 after booster vaccination, intramuscular Ad5-nCoV induced an obvious IgG antibody response, but no IgG antibody response was found in the aerosolized Ad5-nCoV group, indicating that aerosolized Ad5-nCoV stimulated a slower systemic immune response than that stimulated by intramuscular injection. We have not developed an assay to characterize the mucosal immune response, so the mucosal immune response at this time point is unclear. At day 14 after booster vaccination, the systemic immune advantage of aerosolized Ad5-nCoV was fully demonstrated. Only 1/5 of the dose given by intramuscular injection produced a T-cell immune response higher than that of intramuscular Ad5-nCoV. The binding and neutralizing antibodies against the wild-type strain induced by aerosolized Ad5-nCoV were slightly decreased compared to those induced by intramuscular injection, but the level of neutralizing antibodies against the Omicron variant was greater than that induced by intramuscular injection. The neutralizing antibodies from most intramuscular COVID-19 vaccines peak at day 14 after booster vaccination and then decline*(14)*. At day 28 after booster vaccination, the neutralizing antibodies induced by aerosolized Ad5-nCoV still tended to be increased compared to those at day 14, showing different kinetics from other intramuscular booster vaccination regimens examined in this study.

The highly transmissible Omicron variant severely impairs the neutralizing activity of priming two-dose COVID vaccines, with a more than 10-fold reduction compared to that with the wild-type strain*(15-17)*. However, in the present study, the neutralizing activity against Omicron after booster vaccination was ∼2∼3-fold lower than that against the wild-type strain, regardless of which vaccine was used. In fact, a very small number of participants showed no reduction in neutralizing antibodies against the Omicron variant. A similar pattern of neutralization against the Omicron variant was observed in mRNA vaccine before and after booster vaccination*(16, 18, 19)*. An additional booster vaccine dose generated a more potent, cross-reactive antibody responses compared to that induced by the prime vaccination. Repeated antigen exposure improves the affinity maturation of the neutralizing antibodies and increases the potency, breadth and resilience to viral escape mutations of the neutralizing antibodies*(20-22)*.

T-cell immunity is required for viral clearance and supports the generation and maintenance of high-affinity antibodies*(23)*. Adenovirus-vectored COVID-19 vaccines are advantageous as they induce a strong T-cell response, and here, aerosolized Ad5-nCoV at a lower dose induced a more substantial systemic IFNγ cellular response than intramuscular Ad5-nCoV. We speculate that the resident cellular responses are stronger in the airway and lung than in the blood, since a stronger cellular response in the lungs than in the spleen of mice via intranasal vaccination with Ad5-nCoV was observed. Viral mutations have a less pronounced impact on T-cell immunity than on neutralizing antibody responses, which can limit the impact of individual viral mutations and potentially enhance protection against severe disease from SARS-CoV-2 variants.

In the present study, intramuscular or aerosolized Ad5-nCoV-induced neutralizing antibodies and T-cell responses after booster vaccination were significantly higher than those from the recombinant RBD dimer vaccine ZF2001. However, ZF2001 booster vaccination induced 2-fold more neutralizing antibodies than the homologous inactivated vaccine booster. Low cellular responses were detected with the aluminum-adjuvanted recombinant protein and inactivated COVID-19 vaccine. In the case of ChAdOx1 nCoV-19 and BNT162b1-primed vaccination, the immune responses upon booster vaccination with the recombinant protein vaccine (NVX-CoV2373) were also inferior to those upon administration of the adenovirus-vectored boosters, including neutralizing antibodies and cellular immune responses*(10, 24)*. Adenovirus-vectored vaccines are a better alternative to booster regimens based on inactivated vaccine-primed vaccination over recombinant protein vaccines in China.

There are some limitations to this study. First, we have not yet developed appropriate assays for mucosal immune responses to deepen our understanding of the immune advantage of aerosolized Ad5-nCoV. Saliva IgA antibodies were detected in some BNT162b2 vaccine recipients*(25, 26)*, but whether IgA was exuded from serum or produced by a local mucosal immune response could not be determined. More assays need to be developed to analyze local mucosal immune responses, including secretory IgA antibodies and local cellular immune responses. Second, we assessed immunogenicity 28 days after booster vaccination, but further development of the immune response and the longevity of the responses remain to be evaluated. The half-life of serum neutralizing antibodies was 69-173 days for two-dose mRNA vaccine recipients and 103 days for SARS-CoV-2 convalescents*(27-29)*. The long-term antibody dynamics of neutralizing antibodies after the booster vaccination should be further investigated.

In summary, in the face of waning immunity and the circulation of SARS-CoV-2 variants, neutralizing antibody and T-cell responses were boosted most efficiently with aerosolized Ad5-noV in those who received inactivated vaccines as initial doses. Boosters probably increase vaccine effectiveness against infection with and transmission of the SARS-CoV-2 Omicron variant.

## Materials & Methods

### Study design

This was an open and parallel-controlled study, and 904 eligible participants were assigned to four groups to receive one dose of Ad5-nCoV via intramuscular injection (Ad5-nCoV-IM, 5×10^10^ viral particles), aerosolized Ad5-nCoV (Ad5-nCoV-IH, 1×10^10^ viral particles), a recombinant protein subunit vaccine (ZF2001, 25 μg) or an inactivated vaccine (CoronaVac, 3 μg) in December 2021. All participants who received two doses of inactivated vaccine (65.4% receiving CoronaVac, 23.3% receiving BBIBP-CorV (Sinopharm) and 11.3% receiving the mixed CoronaVac and BBIBP-CorV) 6 months prior were enrolled in this study. Participants were excluded if they had known previous COVID-19 infection or had an immunosuppressive condition. Participants were followed longitudinally to evaluate the immune response to different boosters at days 0, 7, 14 and 28 after booster vaccination. The protocol was approved by the Ethics Committee of 305 Hospital of PLA.

### RBD-binding IgG assay

RBD-binding IgG antibodies in the heat-inactivated human serum samples and the culture supernatant of PBMCs stimulated for 4 days with R848 + IL-2 were detected with an RBD-binding IgG ELISA kit (Beijing, Kewei). Briefly, diluted samples and a reference standard were added in duplicate to rSARS-CoV-2 RBD-precoated wells and incubated for 30 min at 37 °C. The microplates were washed, and a horseradish peroxidase (HRP)-conjugated goat anti-human IgG secondary antibody was added to bind the RBD-bound human antibodies. After 30 min of incubation, the microplates were washed, and TMB chromogenic substrate was added to generate a colorimetric signal for 10 min. A stop solution was added to stop color development, and the signal was read on a microplate reader. The total anti-RBD IgG antibody levels were quantitated in ELISA units (EU) ml^-1^ by comparison to a reference standard curve created from monoclonal antibodies against SARS-CoV-2 RBD. The results were analyzed by GraphPad Prism 8.0.2 using 4-PL curve fitting. The WHO international standard for anti-SARS-CoV-2 IgG (NIBSC code 20/136) was used as a reference.

### Pseudotype-based neutralization assays

PNAb assays were performed using the human immunodeficiency virus (HIV) pseudotyped virus production system. HEK293T cells were cotransfected with the plasmids pNL4.3-Luc-R^-^E^-^ and pCAGGS-S^WT^ or pCAGGS-S^Omicron^ with TurboFect transfection reagent (Thermo Scientific). The pCAGGS-S^WT^ and pCAGGS-S^Omicron^ plasmids were constructed and encoded the wild-type (hCoV-19/Wuhan/Hu-1/2019, GISAID EPI_ISL_402125) and Omicron variant (hCoV-19/Botswana/R40B60_BHP_3321001247/2021, GISAID EPI_ISL_6640917) of the SARS-CoV-2 virus spike glycoprotein, respectively. Supernatants were collected 48 hours posttransfection, filtered, aliquoted and frozen at -80 °C before use.

Neutralizing activity in each sample was measured with a serial dilution approach. Each sample was serially diluted 3-fold in duplicate from 1:30 to 1:7290 or 1:21870 in complete DMEM before incubation with the titrated pseudovirus SARS-CoV-2 for 1 hour prior to the addition of 2×10^4^ 293T-ACE2 cells. Following a 48-h incubation period at 37 °C and 5% CO_2_, luciferase activity was determined with the BriteLite Plus Reporter Gene Assay System (PerkinElmer) using a microplate reader (Tecan). EC_50_ neutralization titers were calculated using the Reed-Muench method. The lower limit of detection (LLOD) was 30, and titers below the LLOD were set to 15. The WHO international standard for anti-SARS-CoV-2 IgG (NIBSC code 20/136) was used as a reference.

### Spike-specific IgG ELISpot assays

Spike-specific IgG ELISpot assays were performed on R848- and IL-2-activated PBMCs with a human IgG ELISpot Kit (Mabtech). Briefly, fresh PBMCs were activated with a mixture of R848 at 1 μg ml^-1^ and rhIL-2 at 10 ng ml^-1^ for 4 days. PVDF ELISpot plates were coated with a purified anti-human IgG monoclonal antibody (MT91/145), incubated at 4-8 °C overnight, and blocked with RPMI 1640 medium containing 10% fetal bovine serum and 1× penicillin–streptomycin solution (Gibco) for at least 30 min at room temperature. Activated PBMCs were washed to remove any secreted antibodies, counted, diluted to the indicated concentration, and added to the ELISpot plates. Tests were performed in duplicate, with 20,000-200,000 cells per well in 100 μl of medium. The plates were incubated in a 37 °C humidified incubator with 5% CO_2_ for 16-24 hours. The secretion of spike-specific IgG was visualized by the addition of a biotinylated SARS-CoV-2 spike antigen (1 μg ml^-1^) followed by streptavidin-HRP and TMB substrate. The spots were counted using an ELISpot counter (SinSage Technology), and the results are expressed as SARS-CoV-2 spike-specific IgG spot-forming cells (SFCs) per 10^6^ PBMCs.

### IFNγ ELISpot assays

IFNγ ELISpot assays were performed with fresh PBMCs and a human IFNγ ELISpot Kit (Mabtech) following the manufacturer’s instructions. Tests were performed in duplicate. The precoated ELISpot plates were washed with sterile PBS and blocked with RPMI 1640 medium containing 10% fetal bovine serum and 1× penicillin–streptomycin solution (Gibco) for at least 30 min at room temperature. Fresh PBMCs were added at 2 × 10^5^ cells per well along with the SARS-CoV-2 spike protein peptide pool (1 μg ml^-1^/peptide) or the same volume of DMSO for unstimulated controls. The cells were incubated in a 37 °C humidified incubator with 5% CO_2_ for 16-24 hours. IFNγ spots were detected after the addition of a biotinylated detection antibody (7-B6-1-biotin, 1 μg ml^-1^) followed by streptavidin-HRP and TMB substrate. The spots were counted using an ELISpot counter (SinSage Technology). The counts were summarized as the mean values of duplicate wells with the values of the unstimulated wells subtracted, and negative values were set to zero. The results are expressed as SARS-CoV-2 spike-specific IFNγ SFCs per 10^6^ PBMCs. Responses were considered positive if there were ≥50 spike-specific SFCs per 10^6^ PBMCs and the ratio of spots in the stimulated wells to spots in the background wells was no less than 2.1.

### Statistical analysis

All analyses of participant samples were conducted using GraphPad Prism 8.0.2 or SAS 9.4. Levels of antibodies against SARS-CoV-2 are presented as the GMT or GMC with 95% CI. Spike-specific IgG spots and IFNγ responses are depicted as the median with IQR. Categorical data were analyzed by the χ2 test or Fisher’s exact test. Multiple group comparisons were analyzed by running a nonparametric (Kruskal–Wallis ANOVA) statistical test and corrected using Tukey’s and Dunnett’s tests as indicated in the figure legends. The correlation between concentrations of log-transformed neutralizing antibody and binding antibody levels was analyzed using Spearman’s correlation. P values less than 0.05 were considered to indicate statistical significance.

## Data Availability

All data produced in the present study are available upon reasonable request to the authors

## Acknowledgements

We thank all research participants and staffs in the study.

## Author contributions

Y.F.S. is the principal investigator of this trial. W.C., L.H.H., Y.W.L., and K.L.L. designed the trials and the study protocol. Z.Z., and S.P.W. drafted the manuscript. W.C., and L.L.H. contributed to critical review and revising of the report. Z.Z., S.P.W., Y.W.L., and K.L.L. contributed to data interpretation and revising the manuscript. S.P.W. led laboratory analyses. S.P.W., Z.Z., P.F.F., X.H.S., Y.D.W., Z.H.Z., J.H.X., Y.L., Y.H.L., M.H., G.Y.Z., S.L.L., Y.Z., F.Z.S., Y.L.Y., and X.F.Z. were responsible for laboratory analyses. B.S.W. was responsible for statistical analysis. T.Z., and J.L.Z. contributed to study supervision. X.W.Z., J.S, J.P.Z., K.F.F., P.W.L., P.L., and Z.Y.Q. led and participated in site work, including recruitment, follow-up and data collection.

## Data and materials availability

To protect participants’ confidentiality, the individual participant data that underlie the results reported in this article will only be shared after de-identification. Researchers who provide a scientifically sound proposal will be allowed to access to the de-identified individual participant data. Requests and proposals should be directed to the corresponding author.

